# Factors Associated with Treatment-Resistant Hypertension: Results of a prospective observational Study

**DOI:** 10.1101/2025.05.28.25328513

**Authors:** Mohammed Awais Hameed, Mohamed Elsadig, Shakil Ahmad, M. Sayeed Haque, Charles Ferro, Paramjit Gill, Indranil Dasgupta

## Abstract

**Introduction:** Treatment-resistant hypertension (TRH) is defined as uncontrolled blood pressure despite the use of ≥3 antihypertensive medications at optimum tolerated doses. It is associated with increased risks of cardiovascular events, kidney disease, and mortality. White-coat hypertension, non-adherence, and inappropriate drug combinations overestimate its prevalence. The exact cause of TRH remains unclear, though obesity, obstructive sleep apnoea, and sympathetic overactivity may contribute. This study aimed to better understand the factors associated with true TRH.

**Methods:** Adult patients with treated hypertension without confirmed secondary causes from the West Midlands Hypertension Centre, UK were recruited for comprehensive evaluation. Patients underwent routine clinical assessment, including tests for endothelial function, body composition, and endothelial biomarkers; comparing true TRH with non-TRH patients.

**Results:** Of 141 patients, 60 (43%) had true TRH after excluding whitecoat effect, secondary hypertension and medication non-adherence. The TRH patients were significantly older, had a longer duration of hypertension, and more frequently had diabetes. They had higher rates of left ventricular hypertrophy, higher extracellular water, lower eGFR, and higher urine albumin. They also had higher cardiac biomarkers, (serum NT-proBNP and hs-troponin), inflammatory markers (serum free light chains), aldosterone:renin ratio, and serum Endothelin-1. There was no difference between the groups in adjusted arterial stiffness, reactive hyperaemia or overnight pulse oximetry. Multivariate analysis identified only NT-proBNP as a significant factor associated with TRH (p=0.027).

**Conclusion:** The FACT-RHY study provides valuable insights into the possible pathophysiological mechanisms of TRH. These results emphasize the need for further research into the mechanisms underlying TRH and potential management strategies.

## Introduction

Treatment-resistant hypertension (TRH) is defined as uncontrolled blood pressure (BP) despite the use of at least three antihypertensive medications at maximally tolerated dose including a diuretic. Controlled hypertension on four different antihypertensives is referred to as controlled TRH (1). Regardless of whether it is uncontrolled or controlled, TRH is associated with increased risk of cardiovascular events, kidney failure and death (2, 3).

The reported prevalence of TRH ranges from 5% to 30% (4–9). However, in clinical practice, non-standardised BP measurements, white-coat hypertension, medication non-adherence, non-rational antihypertensive drug combination, or inadequate drug dosing (collectively termed ‘apparent TRH’ or ‘pseudo-resistance’) overestimate the prevalence of TRH (8, 10, 11).

The pathophysiology of TRH remains unclear; although concomitant medications, obstructive sleep apnoea (OSA), obesity and secondary causes of hypertension contribute to treatment resistance. Sympathetic overactivity has been thought to be the underlying mechanism, but the mixed results of sympathetic renal denervation trials till date suggests other mechanisms and factors may be important (12–14). The aim of this study was to explore and understand factors associated with true treatment resistance.

## Methods

A study of factors associated with treatment-resistant hypertension (FACT-RHY) is a prospective, observational study which proposed to comprehensively characterise a group of hypertensive subjects referred to a tertiary hypertension clinic for uncontrolled BP.

Patients aged 18-80 years with treated hypertension were prospectively recruited from the West Midlands Hypertension Centre (WMHC) at Birmingham Heartlands Hospital (May 2017 – July 2021). Patients with white coat-effect using a 24-hour ambulatory BP monitor, an eGFR ≥30 ml/min/1.73m² (Modification of Diet in Renal Disease study [MDRD] equation), renal artery stenosis, pheochromocytoma, primary hyperaldosteronism, Cushing’s disease, and obstructive sleep apnoea (OSA) were excluded. Patients attended a comprehensive evaluation after giving written informed consent.

At the study visit, demographic data including age, sex, and ethnicity were recorded. A detailed history was taken covering the duration of hypertension, medication use (dosage and frequency), smoking habits, and alcohol consumption. Medical history, including angina, myocardial infarction (MI), heart failure, dyslipidaemia, diabetes, stroke, transient ischemic attack (TIA), chronic kidney disease (CKD), and peripheral vascular disease, was reviewed from electronic medical records. Family history of hypertension, cerebrovascular disease, and ischemic heart disease was also documented.

Body mass index (BMI) was calculated from height and weight, and waist and hip measurements were taken to determine waist-to-hip and waist-to-height ratios.

Standardised clinic BP measurement was performed in accordance with the recommendations set out by the British and Irish Hypertension Society (15) using the BpTRU-100 (BpTRU Medical Devices, Coquitlam, BC, Canada). Six BP measurements at 2- minute intervals were taken and the average of last 5 measurements was recorded. A 12- lead ECG was performed, and the Cornell product was calculated to screen for left ventricular hypertrophy (LVH), defined as ≥2440 mm x ms (16).

Whole-body bioimpedance spectroscopy (BIS) was performed using the Fresenius body composition monitor (BCM; Fresenius Medical Care, Bad Homburg, Germany) to assess fluid status in both intracellular and extracellular compartments, with overhydration (OH) quantified as the difference between expected and measured extracellular water. Normal OH ranges from -1.1 to +1.1 L (10th to 90th percentile) in healthy normal population (17).

Pulse wave analysis (PWA) and pulse wave velocity (PWV) were conducted using the Vicorder system (Skidmore Medical, Bristol, UK) to assess arterial stiffness. Augmentation Index, the difference between the first and second systolic peaks of the central BP waveform, is expressed as a percentage of the pulse pressure. PWV was measured over carotid and femoral arteries.

Functional assessment of endothelial function was assessed using EndoPAT 2000 (Itamar Medical, Israel) which measures reactive hyperaemia index (RHI) via peripheral arterial tone. An RHI >1.67 or a log-transformed RHI >0.51 was considered normal.

Blood samples were collected after a 30-minute supine rest and analysed for urea, electrolytes, lipid profile, bone markers, C-reactive protein (CRP), thyroid stimulating hormone (TSH), cortisol, Vitamin D, haemoglobin A1c (HbA1c), renin, aldosterone, troponin I, N-terminal pro B-type natriuretic peptide (NT-proBNP), and serum free light chains (sFLC). Biomarkers for endothelial dysfunction, including soluble fms-like tyrosine kinase-1 (sFLT-1), Endothelin-1 (ET-1), endoglin, Intercellular Adhesion Molecule-1 (ICAM-1), and Vascular Cell Adhesion Molecule-1 (VCAM-1), were measured using enzyme-linked immunosorbent assays (ELISA). Where any parameter values were reported as below the level of quantification, the lower limit of quantification divided by 2 was used to impute a value for any such instance (18, 19).

Urine samples were collected to quantify albumin-to-creatinine ratio (ACR) and test for 23 common antihypertensive drugs or their metabolites. Additionally, 24-hour urine collections were analysed for catecholamines, metadrenalines, and electrolytes, including sodium and potassium, from which daily salt intake was estimated.

Patients completed the Epworth Sleepiness Scale (ESS), which categorizes sleepiness risk as low (≤10) or high (>10) (20). Overnight pulse oximetry was used to screen for OSA, with an oxygen desaturation index (ODI) ≥5 events per hour considered abnormal (21).

Data were recorded in a custom Microsoft Access database and analysed using IBM SPSS Statistics 30 (IBM Corp., Armonk, N.Y., USA). Mean (standard deviation) and median (interquartile range) were reported for parametric and non-parametric data respectively. All data were compared between those with true-TRH and those with non-TRH. Continuous variables were compared using t-tests for parametric data and Mann-Whitney U tests for non-parametric data. Categorical variables were compared with chi-square tests. Multiple linear or logistic regression analysis was performed to assess the impact of independent variables on dependent outcomes. A p-value of <0.05 was considered statistically significant.

Typically, approximately 280 new patients are seen in the WMHC over a year with apparent TRH, young patients with hypertension and patients who multiple intolerances to BP-lowering medications. Around 42% of all new patients meet the criteria for apparent TRH and based on previously published evidence (22, 23), we estimated prevalence of true TRH likely to be 50-60% of those with apparent TRH. Therefore, we expected to find an estimated 60-70 patients with true TRH. The study was approved by the South East Scotland Research Ethics Committee (reference 16/SS/0162) and conducted in accordance with the Declaration of Helsinki and Good Clinical Practice guidelines.

## Results

A total of 141 individuals took part in the study of which 60 (43%, 95% CI: 35% - 51%) were confirmed to have true TRH and remaining 81 were classed as non-TRH in the study. Ninety-two patients (65%, 95% CI: 57% - 73%) were taking three or more antihypertensives of which 60 (65%, 95% CI: 55% - 74%) were classed as TRH based on their BP and adherence to their pharmacological therapy with presence of ≥3 antihypertensives in urine (**Figure 1**). TRH group also comprised of 19 (32%, 95% CI: 21% - 44%) patients prescribed ≥4 antihypertensives with controlled BP (controlled TRH) and 41 patients prescribed ≥3 antihypertensives with uncontrolled BP. Any patient prescribed <3 antihypertensives (n=49) and those prescribed ≥3 antihypertensives found to have pseudo-resistance (n=32) were classified as non-TRH.

**Figure 1:**
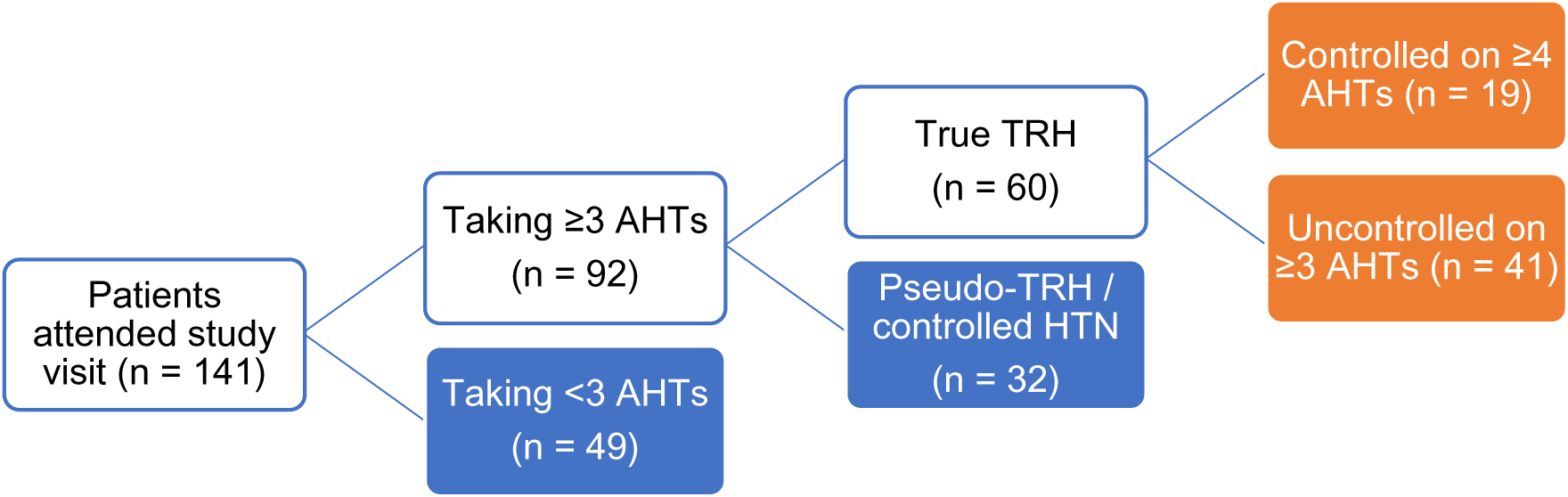
Allocation of patients into groups - treatment-resistant hypertension (orange) and non-TRH (blue). Abbreviations: AHTs, antihypertensives; TRH, treatment-resistant hypertension.

Table 1 shows that TRH patients were significantly older compared to non-TRH patients; mean age 58 years and 46 years respectively (p=0.044). The gender and ethnicity distribution across the two groups was similar. Patients with TRH had a significantly longer duration of hypertension and a higher proportion with diabetes mellitus. There were no significant differences in BP, BMI, waist-to-hip ratio, or waist-to-hip ratio. There was a higher rate of LVH in patients with TRH; 30% compared with 11% in non-TRH group (p= 0.009).

**Table 1:** Baseline demographic and clinical characteristics of all study patients. Data presented as frequencies (percentage), mean ± standard deviation, and median (interquartile range). Log transformed variables are reported as mean (95% confidence interval) and indicated by §.

|  | All Patients | TRH | Non-TRH | P value |
| --- | --- | --- | --- | --- |
| N | 141 | 60 | 81 |  |
| Age years | 51.0 ± 13.9 | 57.8 ± 11.9 | 46.0 ± 13.2 | **0.04** |
| **Sex** |  |  |  | 0.493 |
| Male | 74 | 34 (56.7) | 40 (49.4) |  |
| Female | 67 | 26 (43.3) | 41 (50.6) |  |
| **Ethnicity** |  |  |  | 0.344 |
| White | 77 | 31 (51.7) | 46 (56.8) |  |
| African-Caribbean | 30 | 17 (28.3) | 13 (16.0) |  |
| Asian | 31 | 11 (18.3) | 20 (24.7) |  |
| Mixed | 3 | 1 (1.7) | 2 (2.5) |  |
| Body Mass Index, kg/m<sup>2</sup> | 31.9 ± 5.9 | 32.4 ± 5.2 | 31.5 ± 6.3 | 0.283 |
| Waist to Hip ratio | 0.93 ± 0.11 | 0.96 ± 0.10 | 0.91 ± 0.10 | 0.070 |
| Waist to Height ratio | 0.61 ± 0.09 | 0.63 ± 0.07 | 0.60 ± 0.10 | 0.074 |
| Systolic BP, mmHg<sup>§</sup> | 142 (138 – 146) | 144 (138 – 150) | 141 (136 – 146) | 0.671 |
| Diastolic BP, mmHg | 91 ± 14 | 87 ± 14 | 94 ± 14 | 0.361 |
| Left ventricular hypertrophy | 27 | 18 (30.0) | 9 (11.1) | **0.009** |
| **Smoking Status** |  |  |  | 0.859 |
| Never | 74 | 30 (50.0) | 44 (54.3) |  |
| Ex | 37 | 17 (28.3) | 20 (24.7) |  |
| Current | 30 | 13 (22.0) | 17 (20.3) |  |
| Smoking pack years<sup>§</sup> (n=62) | 16 (5 – 32) | 20 (7 – 47) | 13 (5 – 20) | 0.117 |
| Alcohol intake, units per week (n=38) | 10 ± 6 | 12 ± 7 | 9 ± 5 | 0.124 |
| **Duration of hypertension** |  |  |  | **<0.001** |
| <1-5 years | 47 | 9 (15.0) | 38 (46.9) |  |
| 6-15 years | 48 | 17 (28.3) | 31 (38.3) |  |
| >15 years | 46 | 34 (56.7) | 12 (14.8) |  |
| **Comorbidities** |  |  |  |  |
| Ischaemic heart disease | 16 | 8 (13.3) | 8 (9.9) | 0.710 |
| Diabetes Mellitus | 26 | 17 (28.3) | 9 (11.1) | **0.017** |
| Cerebrovascular disease | 13 | 7 (11.7) | 6 (7.4) | 0.569 |
| Chronic kidney disease | 13 | 9 (15.0) | 4 (4.9) | 0.081 |
| Peripheral vascular disease | 5 | 3 (5.0) | 2 (2.5) | 0.651 |
| **Family History** |  |  |  |  |
| Hypertension | 109 | 42 (70.0) | 67 (82.7) | 0.114 |
| Ischaemic heart disease | 36 | 16 (26.7) | 20 (24.7) | 0.944 |
| Cerebrovascular disease | 35 | 20 (33.3) | 15 (18.5) | 0.069 |

Patients with TRH had a significantly higher number of prescriptions of antihypertensive medications as well as having a higher total pill burden. A significantly higher proportion of patients with TRH were prescribed lipid-lowering therapy; 30 (50%, 95% CI: 38% - 62%) versus 18 (22%, 95% CI: 15% - 32%) in non-TRH group (p=0.001). Virtually all classes of antihypertensive medications were prescribed more frequently for patients with TRH (**table 2**).

**Table 2:** Number of prescribed medications and class of antihypertensives. Data as median (IQR) and counts (percentage is expressed as a proportion of the corresponding column total).

|  | All Patients | TRH | non-TRH | P value |
| --- | --- | --- | --- | --- |
| Number of antihypertensives | 3 (2 – 4) | 4 (3 – 5) | 2 (1 – 3) | <0.001 |
| Total number of medications | 5 (3 – 8) | 7 (5 – 10) | 3 (2 – 6) | <0.001 |
| Lipid lowering therapy | 48 | 30 (50.0) | 18 (22.2) | 0.001 |
| **Class of prescribed antihypertensives** |  |  |  |  |
| Calcium channel blocker | 103 | 51 (85.0) | 52 (64.2) | 0.010 |
| Renin angiotensin aldosterone system inhibitors | 125 | 58 (96.7) | 46 (82.7) | 0.021 |
| Diuretic | 76 | 47 (78.3) | 29 (35.8) | <0.001 |
| Thiazide/thiazide-like | 45 | 27 (45.0) | 18 (22.2) | 0.007 |
| Loop | 24 | 15 (25.0) | 9 (11.1) | 0.052 |
| Potassium-sparing | 29 | 22 (36.7) | 7 (8.6) | <0.001 |
| Beta-adrenergic receptor blocker | 45 | 31 (51.7) | 14 (17.3) | <0.001 |
| Alpha-adrenergic receptor blocker | 41 | 28 (46.7) | 13 (16.0) | <0.001 |
| Others | 13 | 11 (18.3) | 2 (2.5) | 0.003 |

TRH patients had a lower serum potassium 3.9 ± 0.4 mmol/L vs 4.1 ± 0.4 mmol/L (p=0.012), and lower eGFR 74 (59–95) ml/min vs 100 (80–109) ml/min (p<0.001) respectively compared to non-TRH cohort. There were no differences in lipid profile, but the HbA1c was higher in the TRH group. Cardiac biomarkers, NT-proBNP and hs-Troponin, were higher in patients with TRH than non-TRH (p<0.001). The median combined free light chains were also significantly higher in the TRH cohort (**table 3**).

**Table 3:**
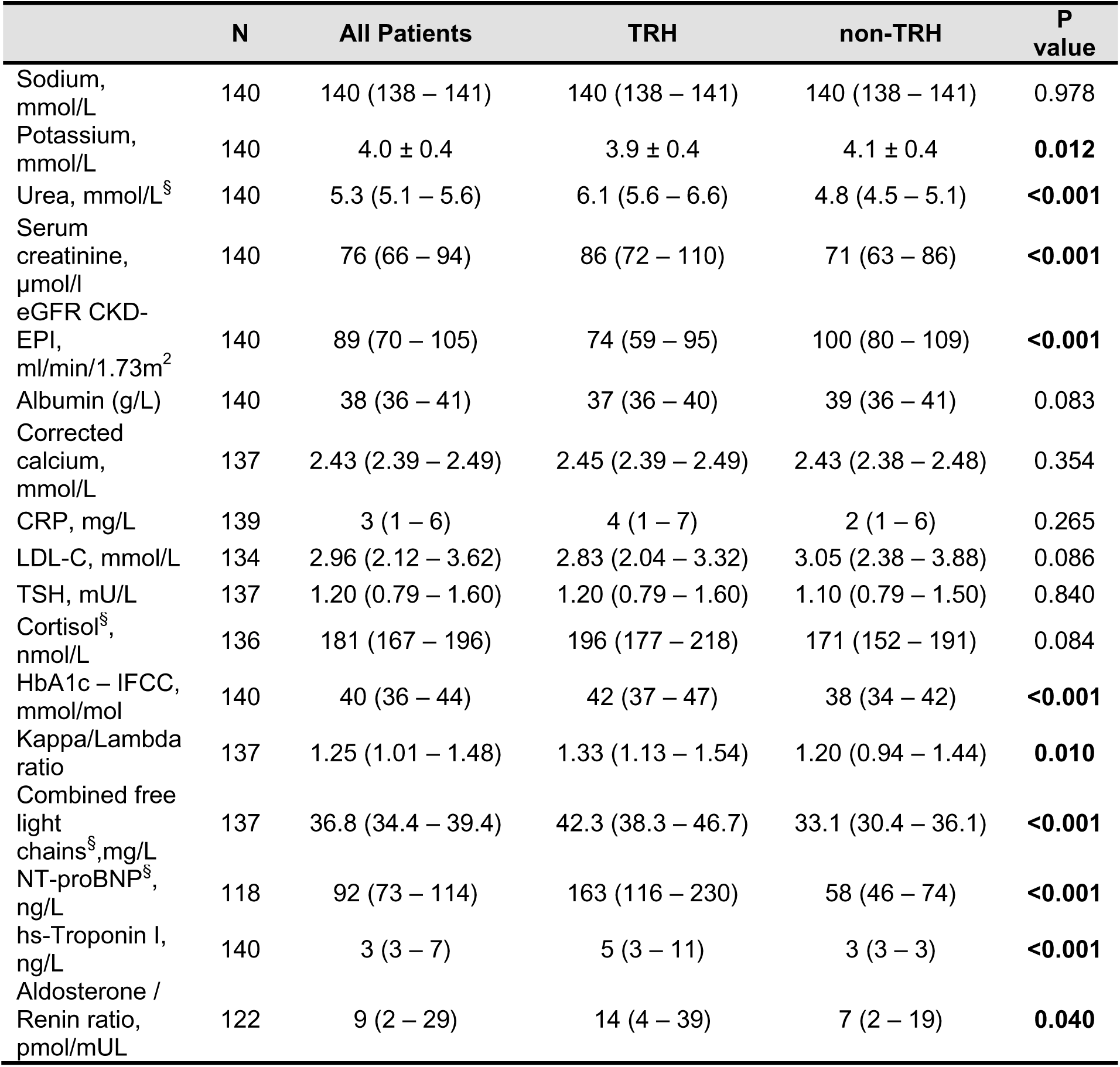
Biochemical characteristics of the study population including TRH and control groups. Data presented as mean ± standard deviation, and median (interquartile range). Log transformed variables are reported as mean (95% confidence interval) and indicated by §.

Median urinary albumin-creatinine ratio was higher in patients with TRH compared to non-TRH; median 2.9 (IQR: 1.1 – 9.8) mg/mmol and median 1.4 (IQR: 0.7 – 7.5) mg/mmol respectively, p=0.036. There were no differences in indices of 24-hour urinary catecholamines and metadrenalines (**table 4**). Daily salt intake as estimated by urinary sodium excretion was comparable between the two groups.

**Table 4:** Spot and 24-hour urine tests in the study population including TRH and control groups.

| Median (IQR) | N | All Patients | TRH | non-TRH | P value |
| --- | --- | --- | --- | --- | --- |
| Albumin / Creatinine ratio, mg/mmol | 137 | 1.6 (0.8 – 8.7) | 2.9 (1.1 – 9.8) | 1.4 (0.7 – 7.5) | **0.036** |
| UF Noradrenaline output, nmol/day | 95 | 271 (161 – 375) | 254 (130 – 399) | 274 (187 – 375) | 0.549 |
| UF Adrenaline output, nmol/day | 94 | 20 (9 – 34) | 16 (7 – 33) | 23 (12 – 38) | 0.155 |
| UF Dopamine output, nmol/day | 97 | 1540<br>(974 – 2359) | 1291<br>(879 – 2241) | 1737<br>(1187 – 2491) | 0.059 |
| Daily salt intake, g/day | 89 | 6.0 (4.4 – 9.1) | 6.1 (4.7 – 10.1) | 6.0 (4.2 – 8.4) | 0.292 |

Both central and peripheral median systolic and pulse pressures were higher in the TRH group, (**table 5**). Augmentation index (both unadjusted and adjusted to a heart rate of 75 bpm) did not differ significantly between groups. Median carotid-femoral PWV (cfPWV) was significantly higher in TRH patients than controls; 9.8 (IQR: 9.3 – 10.3) m/s vs. 8.9 (IQR: 8.5 – 9.3) m/s (p = 0.008). Stroke volume was also higher in the TRH group, but other haemodynamic parameters did not differ significantly.

**Table 5:** Haemodynamic parameters of arterial pulse wave analysis in the entire study population stratified according to patient groups. Log transformed variables are reported as mean (95% confidence interval) and indicated by §.

|  | All Patients | TRH | non-TRH | P value |
| --- | --- | --- | --- | --- |
| cfPWV<sup>§</sup>, m/s | 9.3 (9.0 – 9.6) | 9.8 (9.3 – 10.3) | 8.9 (8.5 – 9.3) | **0.008** |
| Alx, % | 26.9 ± 8.7 | 28.1 ± 7.7 | 26.0 ± 9.3 | 0.167 |
| Alx<sub>75</sub>, (%) | 24.5<br>(20.8 – 31.4) | 25.3<br>(21.2 – 31.7) | 24.3<br>(20.3 – 31.0) | 0.438 |
| Heart rate, bpm | 66 ± 12 | 61 ± 11 | 70 ± 12 | **<0.001** |
| Peripheral SBP, mmHg | 169 (154 – 187) | 162 (147 – 172) | 151 (137 – 164) | **0.010** |
| Peripheral DBP, mmHg | 81 (75 – 90) | 81 (73 – 88) | 81 (75 – 90) | 0.346 |
| Peripheral PP, mmHg | 73 (60 – 85) | 78 (67 – 90) | 68 (58 – 78) | **<0.001** |
| Central SBP<sup>§</sup>, mmHg | 152 (148 – 156) | 156 (151 – 162) | 149 (144 – 153) | **0.027** |
| Central PP<sup>§</sup>, mmHg | 69 (66 – 72) | 75 (71 – 80) | 65 (62 – 69) | **<0.001** |
| MAP<sup>§</sup>, mmHg | 112 (109 – 114) | 112 (108 – 115) | 112 (109 – 115) | 0.999 |
| Cardiac output, L/min | 6.9 (6.0 – 8.6) | 6.9 (6.0 – 8.7) | 6.9 (6.0 – 8.4) | 0.702 |
| Total peripheral resistance, mmHg.min.mL<sup>-1</sup> | 0.98 (0.83 – 1.09) | 0.97 (0.82 – 1.09) | 0.99 (0.84 – 1.09) | 0.852 |
| Stroke volume<sup>§</sup>, mL | 109 (104 – 114) | 118 (110 – 127) | 103 (96 – 109) | **0.004** |

Multiple regression (supplementary table 3) assessed whether the association between TRH and cfPWV remained after adjusting for age, MAP, and HR. The final model explained 49.0% of variance in cfPWV, F(4,133) = 31.912, p<0.001. TRH was added in stepwise manner to the regression model and was found to be nonsignificant (supplementary table 3).

Overhydration derived from the BCM is presented as absolute values in litres and as a percentage of ECW. The mean absolute OH and percentage OH were significantly higher in the TRH compared to control (supplementary table 4). 44% of all study participants were found to be underhydrated with an absolute OH less than -1.1 litres with the remaining 51% normally hydrated.

The mean ECW-to-ICW ratio was higher in the TRH group compared to controls; 0.87 ± 0.09) and 0.82 ± 0.08 respectively (p<0.001). Similarly, mean ECW-to-TBW ratio was higher in the TRH group comparatively; 0.47 ± 0.03 and 0.45 ± 0.02 respectively (p<0.001). However, the magnitude of the mean difference between the two groups was small.

The results of biomarkers of endothelial dysfunction and EndoPAT evaluation are summarised in **table 6**. Endothelial function as assessed by EndoPAT was carried out in half of the total study population (n=71, 50%). There was no difference in the RHI in the two groups of patients. Similarly, when patients were divided into normal or abnormal groups based on the LnRHI, there was no difference in the proportion of patients with abnormal endothelial dysfunction in patients with TRH compared to controls; 10 (32.3%) vs 9 (22.5%) respectively (p=0.515).

**Table 6:** Assessment of endothelial dysfunction: EndoPAT measured reactive hyperaemia index and ELISA measured biomarkers.

|  | N | All Patients | TRH | non-TRH | P value |
| --- | --- | --- | --- | --- | --- |
| Reactive hyperaemia index | 71 | 1.97<br>(1.65 – 2.40) | 1.88<br>(1.61 – 2.27) | 2.00<br>(1.80 – 2.48) | 0.278 |
| LnRHI | 71 | 0.70 ± 0.24 | 0.66 ± 0.21 | 0.74 ± 0.26 | 0.204 |
| EndoPAT<br>Endothelial function |  |  |  |  | 0.515 |
| Normal |  | 52 (73.2) | 21 (67.7) | 31 (77.5) |  |
| Abnormal |  | 19 (26.8) | 10 (32.3) | 9 (22.5) |  |
| sFLT-1, pg/mL | 141 | 297.07<br>(254.28 – 359.60) | 299.26<br>(264.52 – 364.91) | 297.07<br>(250.70 – 349.34) | 0.585 |
| Endoglin, ng/mL | 140 | 4.69 (3.91 – 5.34) | 4.52 (3.74 – 5.32) | 4.78 (4.12 – 5.38) | 0.196 |
| Endothelin-1, pg/mL | 141 | 2.16<br>(1.71 – 2.59) | 2.24<br>(1.84 – 2.85) | 2.03<br>(1.55 – 2.40) | **0.010** |
| VCAM-1, ng/mL | 141 | 1009.9<br>(761.7 – 1141.3) | 1164.1<br>(769.2 – 1190.8) | 993.3<br>(741.3 – 1087.5) | 0.067 |
| ICAM-1, ng/mL | 137 | 105.80<br>(86.41 – 150.54) | 110.41<br>(89.30 – 168.45) | 104.16<br>(85.86 – 144.68) | 0.488 |

Of the soluble endothelial markers, only median endothelin-1 was found to be higher in patients with TRH compared to control group of patients; 2.24 (1.84 – 2.85) pg/mL and 2.03 (1.55 – 2.40) pg/mL (p=0.01). There was no difference in serum concentrations of sFLT-1, endoglin, VCAM-1 and ICAM-1 in the two groups of patients.

Median Eppworth sleepiness score was similar across the two study groups and no siginifcant differences were detected when patients were categorised into low-risk (ESS ≤ 10) or high-risk groups (ESS >10). Seventy-Eight patients (55%) completed overnight pulse oximetry of which 68 completed the minimum 4 hours of recording. None of the parameters (oxygen desaturation index, nocturnal nadir oxygen saturation and proportion of time with oxygen saturation <90%) was significantly different among the study groups (Supplementary table 5).

A multivariable binary logistic regression analysis was performed to assess if any of the biologically plausible significant factors identified is independently associated with TRH. The model included age, sex, average systolic blood pressure, presence of diabetes, cfPWV, OH as percentage of ECW, ARR, cFLC, endothelin-1, hs-troponin I, eGFR and NT-proBNP. The analysis included 97 patients; only NT-proBNP was found to be associated with TRH (p=0.027).

## Discussion

Our study is the largest observational study to date that describes the demographic, biochemical and physiological characteristics of patients with true TRH. Patients with true TRH were older, with longer duration of hypertension, and had a higher prevalence of diabetes mellitus and LVH when compared to a control group of non-TRH patients. The laboratory results indicate that patients with TRH had higher serum creatinine, aldosterone/renin ratio, HbA1c, kappa/lambda ratio, NT-proBNP, urine ACR, and troponin I, as well as lower serum potassium and eGFR compared to non-TRH patients. Although there was no difference between the groups in flow mediated vasodilation as a marker of endothelial function, serum endothelin concentration was higher in the TRH group. On multivariate analysis only NT-proBNP was found to be associated with TRH.

Many of these findings are not unexpected, given that TRH typically occurs in older individuals with a long history of hypertension. The presence of higher ACR and lower eGFR are anticipated, as both hypertension and diabetes mellitus are major contributors to renal albuminuria renal impairment.

In the FACT-RHY cohort, patients with true TRH exhibited significantly higher plasma aldosterone/renin ratio (ARR), a finding consistent with previous studies (24, 25). These patients also had significantly lower serum potassium levels compared to those without TRH, although the absolute difference between the two groups was small. While a low serum potassium level is often considered an early indicator of hyperaldosteronism, recent evidence suggests that only a minority of patients with this condition present with hypokalaemia (26, 27).

Recent studies have highlighted the effectiveness of pharmacological agents that target aldosterone antagonism and synthesis in achieving blood pressure control among patients with TRH (13, 28, 29). Our findings align with these studies, demonstrating that patients with TRH exhibit elevated aldosterone levels and may therefore benefit from such therapeutic interventions.

ARR is the most used screening tool for hyperaldosteronism, but both plasma renin activity and aldosterone concentrations can be influenced by use of antihypertensive medications, dietary sodium intake, and serum potassium levels, which may skew the results in either direction (30). In this study, the higher use of thiazide and loop diuertics may also explain the slightly lower serum potassium in the TRH patients.

In our study, patients with TRH had significantly higher levels of cardiac biomarkers, serum troponin-I and NT-proBNP, which may indicate underlying left ventricular hypertrophy or heart failure. However, none of the patients in our study had a confirmed history of heart failure. Very few patients had prior echocardiography to assess cardiac structure or function, which could provide relevance of these findings. Importantly, NT-proBNP is independently associated with TRH. In hypertensive population, NT-proBNP independently predicts increased risk of incident cardiovascular disease and all-cause mortality across a range of BP categories (31). This may, at least in part, explain the higher risk of cardiovascular events and mortality associated with TRH compared with non-TRH. A strategy of titrating BP-lowering medication especially diuretics based on NT-proBNP could be considered in future studies.

Free light chains (FLCs), markers of B cell activation, are elevated in a range of inflammatory conditions (32). Combined FLCs (cFLCs) have been identified as an independent risk factor for mortality in the general population (33, 34), as well as for mortality and progression to kidney failure in patients with CKD (35), and mortality in those with acute decompensated heart failure (36). In the FACT-RHY study, we found, for the first time, that patients with TRH had significantly elevated cFLCs and kappa to lambda FLC ratio. The cause of elevated serum cFLCs in hypertension remains unclear, although one hypothesis suggests that increased immunoglobulin production may result from vascular damage caused by high BP (37).

Alternatively, the elevated cFLCs may reflect a generalized immune system activation or inflammation. Additionally, since FLCs can be elevated in renal impairment, their levels may also be influenced by decreased renal clearance, which may be a factor in some TRH patients in the FACT-RHY cohort.

This study found no significant difference in cfPWV between patients with true TRH from those with non-TRH, after adjusting for age, MAP and HR. Arterial stiffness is a well-established risk factor for cardiovascular diseases including MI, heart failure, and mortality as well as stroke, dementia and CKD (38–46). Although several studies have explored arterial stiffness in TRH, but results have been inconsistent (47–51).

One possible explanation for the lack of difference in the arterial stiffness between TRH and non-TRH patients could be the higher use of antihypertensive medications in the TRH group.

These medications, including diuretics, calcium channel blockers (CCBs), nitrates, and RAAS inhibitors, are known to reduce arterial stiffness (52). Diuretics and CCBs have been shown to improve outcomes in isolated systolic hypertension, a common feature of aging and arterial stiffness (53–55). However, there is a lack of large randomized controlled trials (RCTs) designed specifically to assess the effect of antihypertensive medications on arterial stiffness and subsequent cardiovascular outcomes, though such a trial is currently underway (56).

Notably, most antihypertensive medications primarily address the dynamic vasoconstrictive component of arterial stiffness and do not target the underlying structural and vascular signalling changes that contribute to arterial stiffening (52).

The BIS analysis in this study found that patients with true TRH had significant expansion in ECW compared to patients without TRH. While ECW expansion contributes to hypertension (57), demonstrating excess ECW is challenging. BIS is commonly used in patients with kidney failure on dialysis to monitor fluid status and guide therapy. In the FACT-RHY study, BIS was employed to explore the relationship between body composition and TRH. While not routinely used in essential hypertension, BIS may be beneficial in cases of difficult-to-control hypertension or TRH (58, 59). Impedance cardiography, which measures hemodynamic profiles and thoracic fluid content to guide diuretic therapy, shares principles with BIS. A meta-analysis showed that impedance cardiography helped patients achieve better BP control (59) and is cost-effective (60). In the FACT-RHY study, whole-body fluid content was estimated with BIS, while impedance cardiography uses segmental BIS to calculate thoracic fluid content.

ELISA analysis revealed significantly higher levels of serum ET-1 in patients with TRH, indicating endothelial dysfunction compared to non-TRH patients. These results suggest that ET-1-induced vasoconstriction may contribute to TRH. Clinical trials of the ETA receptor antagonist darusentan and aprocitentan support this, showing their effectiveness in lowering BP in TRH patients (14, 61–63). Additionally, darusentan has been shown to reduce vascular hypertrophy, indicating a direct role of ET-1 in vascular growth (64, 65). Endothelin antagonists may herald a new era in the management of TRH (66).

Interestingly, functional assessment of endothelial dysfunction using PAT did not differ significantly between the two groups, which is not consistent with the biomarker findings. This discrepancy can be explained by the different mechanisms involved: while ET-1 causes local vasoconstriction, PAT measures nitric oxide-mediated vasodilation in response to reactive hyperaemia. As such, impairment in one pathway may not necessarily affect the other.

## Strengths and limitations

The FACT-RHY study’s strength lies in its prospective design with a cohort of patients from a specialist hypertension clinic. Uniquely, all patients underwent objective adherence testing using urine AHS which, along with the use of ABPM at the first clinic visit to rule out whitecoat effect, systematically excluded apparent or ‘pseudo’ TRH. ABPM can be uncomfortable and was not repeated within the study to minimize patient burden. BpTRU was used at the study visit to further reduce whitecoat effect and ensure accurate classification of patients. Additionally, patients with a history of secondary hypertension were excluded, and all underwent comprehensive screening for secondary causes. This rigorous approach means that the TRH group in the FACT-RHY study represents a true resistant cohort, making the findings more reliable.

The main limitation is it is observational, which prevents establishing causality or measuring the role of specific factors in the development of TRH. A prospective cohort study with matched controls and repeated assessments would be ideal but would require significant resources and time due to the low prevalence of true TRH and the long duration of hypertension before treatment resistance occurs. Despite this, the findings are consistent with other experimental and observational studies in both animal models and humans (14, 50, 51, 65, 67). Another limitation is the small sample size of the TRH cohort. While the target was to recruit up to 140 patients, fewer true TRH patients were identified than expected after ruling out whitecoat effect and nonadherence to medication. Additionally, less than a quarter of those screened and fewer than a third of eligible participants consented to participate in the study. The recruitment challenges were likely due to conducting the study during the COVID-19 pandemic and perceived lack of direct benefit for taking part in this observational study. Furthermore, the study’s recruitment from a specialist hypertension clinic might have introduced selection bias, limiting the generalizability of the findings. Specialist clinics typically see patients with difficult-to-treat hypertension or suspected secondary hypertension, and selecting a control group from this population may not represent the broader hypertensive population. Conversely, recruiting from the community would require a much larger screening effort, and would logistically be challenging.

The results of FACT-RHY study have indicated possible role of a multitude of pathophysiological mechanisms in TRH. Further research is warranted to confirm these finding in larger cohorts and to explore therapeutic strategies based on the findings.

## Data Availability

Data will be available on request with appropriate requests.

## Acknowledgment

The study has formed part of PhD for MAH and ME. Funded by Renal Research Fund, Heartlands Hospital, Birmingham, UK.

## Disclosures

Authors have no conflicts of interest to declare.

## Notes

### Competing Interest Statement

The authors have declared no competing interest.

### Clinical Trial

Not applicable

### Funding Statement

Non-commercial funding.

### Author Declarations

The study was approved by the South East Scotland Research Ethics Committee (reference 16/SS/0162) and conducted in accordance with the Declaration of Helsinki and Good Clinical Practice guidelines.

## References

1. Whelton PK, Carey RM, Aronow WS, et al. 2017 ACC/AHA/AAPA/ABC/ACPM/AGS/APhA/ASH/ASPC/NMA/PCNA Guideline for the prevention, detection, evaluation, and management of high blood pressure in adults: a report of the American College of Cardiology/American Heart Association task force on clinical practice guidelines. J Am Coll Cardiol. 2018;71(19):e127–e248.

2. Daugherty SL, Powers JD, Magid DJ, et al. Incidence and prognosis of resistant hypertension in hypertensive patients. Circulation. 2012;125(13):1635–42.

3. Irvin MR, Booth JN, Shimbo D, et al. Apparent treatment-resistant hypertension and risk for stroke, coronary heart disease, and all-cause mortality. Journal of the American Society of Hypertension. 2014;8(6):405–13.

4. Hajjar I, Kotchen TA. Trends in prevalence, awareness, treatment, and control of hypertension in the united states, 1988-2000. JAMA. 2003;290(2):199–206.

5. Cushman WC, Ford CE, Cutler JA, et al. Success and predictors of blood pressure control in diverse North American settings: the Antihypertensive and Lipid-Lowering treatment to prevent Heart Attack Trial (ALLHAT). The Journal of Clinical Hypertension. 2002;4(6):393–404.

6. Egan BM, Zhao Y, Axon RN, et al. Uncontrolled and apparent treatment resistant hypertension in the United States, 1988 to 2008. Circulation. 2011;124(9):1046–58.

7. Persell SD. Prevalence of resistant hypertension in the United States, 2003–2008. Hypertension. 2011;57(6):1076–80.

8. de la Sierra A, Segura J, Banegas JR, et al. Clinical features of 8295 patients with resistant hypertension classified on the basis of ambulatory blood pressure monitoring. Hypertension. 2011;57(5):898–902.

9. Hanselin MR, Saseen JJ, Allen RR, et al. Description of antihypertensive use in patients with resistant hypertension prescribed four or more agents. Hypertension. 2011;58(6):1008–13.

10. Hayek SS, Abdou MH, Demoss BD, et al. Prevalence of resistant hypertension and eligibility for catheter-based renal denervation in hypertensive outpatients. American Journal of Hypertension. 2013;26(12):1452–8.

11. Hayes P, Casey M, Glynn LG, et al. Prevalence of treatment-resistant hypertension after considering pseudo-resistance and morbidity: a cross-sectional study in Irish primary care. British Journal of General Practice. 2018;68(671):e394–e400.

12. Fadl Elmula FEM, Hoffmann P, Larstorp AC, et al. Adjusted drug treatment is superior to renal sympathetic denervation in patients with true treatment-resistant hypertension. Hypertension. 2014;63(5):991–9.

13. Freeman MW, Halvorsen YD, Marshall W, et al. Phase 2 Trial of Baxdrostat for Treatment-Resistant Hypertension. N Engl J Med. 2023;388(5):395–405.

14. Schlaich MP, Bellet M, Weber MA, et al. Dual endothelin antagonist aprocitentan for resistant hypertension (PRECISION): a multicentre, blinded, randomised, parallel-group, phase 3 trial. The Lancet. 2022;400(10367):1927–37.

15. British and Irish Hypertension Society. Blood pressure measurement: using automated blood pressure monitors 2017 [Available from: https://bihsoc.org/wp-content/uploads/2017/11/BP-Measurement-Poster-Automated-2017.pdf.

16. Okin PM, Roman MJ, Devereux RB, et al. Electrocardiographic identification of increased left ventricular mass by simple voltage-duration products. Journal of the American College of Cardiology. 1995;25(2):417–23.

17. Passauer J, Petrov H, Schleser A, et al. Evaluation of clinical dry weight assessment in haemodialysis patients using bioimpedance spectroscopy: a cross-sectional study. Nephrology Dialysis Transplantation. 2009;25(2):545–51.

18. Senn S, Holford N, Hockey H. The ghosts of departed quantities: approaches to dealing with observations below the limit of quantitation. Stat Med. 2012;31(30):4280–95.

19. Keizer RJ, Jansen RS, Rosing H, et al. Incorporation of concentration data below the limit of quantification in population pharmacokinetic analyses. Pharmacol Res Perspect. 2015;3(2):e00131.

20. Johns MW. A new method for measuring daytime sleepiness: the Epworth Sleepiness Scale. Sleep. 1991;14(6):540–5.

21. Ling IT, James AL, Hillman DR. Interrelationships between body mass, oxygen desaturation, and apnea-hypopnea indices in a sleep clinic population. Sleep. 2012;35(1):89–96.

22. Lawson AJ, Hameed MA, Brown R, et al. Nonadherence to antihypertensive medications is related to pill burden in apparent treatment-resistant hypertensive individuals. Journal of Hypertension. 2020;38(6):1165–73.

23. Thomas O, Shipman KE, Day K, et al. Prevalence and determinants of white coat effect in a large UK hypertension clinic population. Journal Of Human Hypertension. 2015;30:386.

24. Gaddam KK, Nishizaka MK, Pratt-Ubunama MN, et al. Characterization of resistant hypertension: association between resistant hypertension, aldosterone, and persistent intravascular volume expansion. Arch Intern Med. 2008;168(11):1159–64.

25. Calhoun David A, Nishizaka Mari K, Zaman Mohammad A, et al. Hyperaldosteronism among Black and White subjects with resistant hypertension. Hypertension. 2002;40(6):892–6.

26. Rossi GP, Bernini G, Caliumi C, et al. A prospective study of the prevalence of primary aldosteronism in 1,125 hypertensive patients. Journal of the American College of Cardiology. 2006;48(11):2293–300.

27. Mulatero P, Stowasser M, Loh K-C, et al. Increased diagnosis of primary aldosteronism, including surgically correctable forms, in centers from five continents. The Journal of Clinical Endocrinology & Metabolism. 2004;89(3):1045–50.

28. Laffin LJ, Kopjar B, Melgaard C, et al. Lorundrostat Efficacy and Safety in Patients with Uncontrolled Hypertension. 2025;392(18):1813–23.

29. Williams B, MacDonald TM, Morant S, et al. Spironolactone versus placebo, bisoprolol, and doxazosin to determine the optimal treatment for drug-resistant hypertension (PATHWAY-2): a randomised, double-blind, crossover trial. The Lancet. 2015.

30. Funder JW, Carey RM, Mantero F, et al. The management of primary aldosteronism: case detection, diagnosis, and treatment: an Endocrine Society clinical practice guideline. The Journal of Clinical Endocrinology & Metabolism. 2016;101(5):1889–916.

31. Hussain A, Sun W, Deswal A, et al. Association of NT-ProBNP, Blood Pressure, and Cardiovascular Events: The ARIC Study. Journal of the American College of Cardiology. 2021;77(5):559–71.

32. Brebner JA, Stockley RA. Polyclonal free light chains: a biomarker of inflammatory disease or treatment target? F1000 medicine reports. 2013;5(4).

33. Anandram S, Assi LK, Lovatt T, et al. Elevated, combined serum free light chain levels and increased mortality: a 5-year follow-up, UK study. Journal of clinical pathology. 2012;65(11):1036–42.

34. Dispenzieri A, Katzmann JA, Kyle RA, et al. Use of nonclonal serum immunoglobulin free light chains to predict overall survival in the general population. Mayo Clinic proceedings. 2012;87(6):517–23.

35. Ritchie J, Assi LK, Burmeister A, et al. Association of serum Ig free light chains with mortality and ESRD among patients with nondialysis-dependent CKD. Clinical Journal of the American Society of Nephrology. 2015;10(5):740–9.

36. Shantsila E, Wrigley B, Lip GYH. Free light chains in patients with acute heart failure secondary to atherosclerotic coronary artery disease. The American Journal of Cardiology. 2014;114(8):1243–8.

37. Kristensen BØ, Sølling K. Serum concentrations of immunoglobulins and free light chains before and after vascular events in essential hypertension. Acta Medica Scandinavica. 1983;213(1):15–20.

38. Blacher J, Asmar R, Djane S, et al. Aortic pulse wave velocity as a marker of cardiovascular risk in hypertensive patients. Hypertension. 1999;33(5):1111–7.

39. Chae CU, Pfeffer MA, Glynn RJ, et al. Increased pulse pressure and risk of heart failure in the elderly. JAMA. 1999;281(7):634–43.

40. Franklin SS, Larson MG, Khan SA, et al. Does the relation of blood pressure to coronary heart disease risk change with aging? The Framingham Heart Study. Circulation. 2001;103(9):1245–9.

41. Mitchell GF, Moyé LA, Braunwald E, et al. Sphygmomanometrically determined pulse pressure is a powerful independent predictor of recurrent events after myocardial infarction in patients with impaired left ventricular function. Circulation. 1997;96(12):4254–60.

42. Vaccarino V, Berger AK, Abramson J, et al. Pulse pressure and risk of cardiovascular events in the systolic hypertension in the elderly program. The American Journal of Cardiology. 2001;88(9):980–6.

43. Kostis JB, Lawrence-Nelson J, Ranjan R, et al. Association of increased pulse pressure with the development of heart failure in SHEP. American Journal of Hypertension. 2001;14(8):798–803.

44. Blacher J, Guerin AP, Pannier B, et al. Impact of aortic stiffness on survival in end-stage renal disease. Circulation. 1999;99(18):2434–9.

45. Forette F, Seux M-L, Staessen JA, et al. Prevention of dementia in randomised double-blind placebo-controlled Systolic Hypertension in Europe (Syst-Eur) trial. The Lancet. 1998;352(9137):1347–51.

46. Benetos A, Rudnichi A, Safar M, et al. Pulse pressure and cardiovascular mortality in normotensive and hypertensive subjects. Hypertension. 1998;32(3):560–4.

47. Dudenbostel T, Acelajado MC, Pisoni R, et al. Refractory hypertension: evidence of heightened sympathetic activity as a cause of antihypertensive treatment failure. Hypertension. 2015;66(1):126–33.

48. Barbaro NR, Fontana V, Modolo R, et al. Increased arterial stiffness in resistant hypertension is associated with inflammatory biomarkers. Blood Press. 2015;24(1):7–13.

49. Figueiredo VN, Yugar-Toledo JC, Martins LC, et al. Vascular stiffness and endothelial dysfunction: correlations at different levels of blood pressure. Blood pressure. 2012;21(1):31–8.

50. Brandt MC, Reda S, Mahfoud F, et al. Effects of renal sympathetic denervation on arterial stiffness and central hemodynamics in patients with resistant hypertension. Journal of the American College of Cardiology. 2012;60(19):1956–65.

51. Sabbatini AR, Barbaro NR, de Faria AP, et al. Increased circulating tissue inhibitor of metalloproteinase-2 is associated with resistant hypertension. The Journal of Clinical Hypertension. 2016;18(10):969–75.

52. Zieman SJ, Melenovsky V, Kass DA. Mechanisms, pathophysiology, and therapy of arterial stiffness. Arteriosclerosis, Thrombosis, and Vascular Biology. 2005;25(5):932–43.

53. Cushman WC, Materson BJ, Williams DW, et al. Pulse pressure changes with six classes of antihypertensive agents in a randomized, controlled trial. Hypertension. 2001;38(4):953–7.

54. Staessen JA, Fagard R, Thijs L, et al. Randomised double-blind comparison of placebo and active treatment for older patients with isolated systolic hypertension. The Lancet. 1997;350(9080):757–64.

55. SHEP Cooperative Research Group. Prevention of stroke by antihypertensive drug treatment in older persons with isolated systolic hypertension: final results of the Systolic Hypertension in the Elderly Program (SHEP). JAMA. 1991;265(24):3255–64.

56. Sharman JE, Boutouyrie P, Laurent S. Arterial (aortic) stiffness in patients with resistant hypertension: from assessment to treatment. J Current Hypertension Reports. 2017;19(1):2.

57. Guyton AC, Manning RD, Jr., Hall JE, et al. The pathogenic role of the kidney. Journal of cardiovascular pharmacology. 1984;6 Suppl 1:S151–61.

58. Covic A, Voroneanu L, Goldsmith D. Routine bioimpedance-derived volume assessment for all hypertensives: a new paradigm. American Journal of Nephrology. 2014;40(5):434–40.

59. Ferrario CM, Flack JM, Strobeck JE, et al. Individualizing hypertension treatment with impedance cardiography: a meta-analysis of published trials. Ther Adv Cardiovasc Dis. 2010;4(1):5–16.

60. Ferrario CM, Smith RD, for the CONTROL Trial Investigators. Cost-effectiveness of impedance cardiography testing in uncontrolled hypertension. American Heart Hospital Journal. 2006;4(4):279–89.

61. Black HR, Bakris GL, Weber MA, et al. Efficacy and safety of darusentan in patients with resistant hypertension: results from a randomized, double-blind, placebo-controlled dose-ranging study. J Clin Hypertens (Greenwich). 2007;9(10):760–9.

62. Bakris GL, Lindholm LH, Black HR, et al. Divergent results using clinic and ambulatory blood pressures: report of a darusentan-resistant hypertension trial. Hypertension. 2010;56(5):824–30.

63. Weber MA, Black H, Bakris G, et al. A selective endothelin-receptor antagonist to reduce blood pressure in patients with treatment-resistant hypertension: a randomised, double-blind, placebo-controlled trial. The Lancet. 2009;374(9699):1423-31.

64. Park JB, Schiffrin EL. ET(A) receptor antagonist prevents blood pressure elevation and vascular remodeling in aldosterone-infused rats. Hypertension. 2001;37(6):1444–9.

65. Schiffrin EL, Larivière R, Li JS, et al. Enhanced expression of the endothelin-1 gene in blood vessels of DOCA-salt hypertensive rats: correlation with vascular structure. Journal of Vascular Research. 1996;33(3):235–48.

66. Chapman GB, Dhaun N. Endothelin Antagonism: A New Era for Resistant Hypertension? 2025;82(4):611–4.

67. Murray CJ. Increased plasma endothelin level in patients with essential hypertension. N Engl J Med. 1990;322(3):205-.

